# Medical Multimorbidity in Patients with Treatment-Resistant Psychosis and Rare Copy Number Variants: A Retrospective Case Series of 24 Patients

**DOI:** 10.1101/2025.05.13.25325400

**Authors:** Tyler E. Dietterich, Rose Mary Xavier, Maya L. Lichtenstein, Matthew K. Harner, Lisa Bruno, Robert Stowe, Martilias Farrell, Rita A. Shaughnessy, Jonathan S. Berg, Patrick F. Sullivan, Richard C. Josiassen

## Abstract

Neurodevelopmental disorder-risk CNVs (NDD CNVs) are associated with complex neuropsychiatric phenotypes. These CNVs also confer risk for a host of medical outcomes in adults yet the long-term health consequences in the context of comorbid psychiatric illness have not been well documented. Twenty-four psychiatric inpatients with treatment-resistant psychosis were identified as carriers of NDD CNVs as part of a larger Pennsylvania State Hospital genomics study. Comprehensive life course phenotyping was performed through review of medical records, specialized neurobehavioral evaluation, and synthesis of data using the Human Phenotype Ontology. Phenotypes were examined across the cohort and within sets of individuals with CNVs in common. Retrospective phenotyping indicated comorbid medical manifestations across multiple organ systems. Cardiovascular disorders were present in 96% of cases and motor disorders in 92%. All cases had multiple organ system involvement, and most organ systems (12/17 systems) were affected in 50% or more of cases culminating in a high degree of individual-level comorbidity. Potentially novel health outcomes are described for individual CNV loci. Our descriptive case series supports a complex and multidimensional course of illness. Thorough reporting on the long-term implications of these variants is the first step toward advancing clinical care for these complex psychiatric patients carrying NDD CNVs.

## Introduction

People with schizophrenia suffer disproportionately from chronic diseases including cardiovascular diseases, non-insulin-dependent diabetes mellitus, and pulmonary diseases (Brink et al., 2019; Jeste, Gladsjo, Lindamer, & Lacro, 1996; Laursen, Munk-Olsen, & Gasse, 2011). This excess medical comorbidity is a major contributor to the overall suffering, disability, health care utilization, and increased mortality rates in this population (Brink et al., 2019; Launders et al., 2022; Laursen et al., 2011). Poorer health outcomes in schizophrenia are attributed to a number of factors including lifestyle habits (i.e. increased rates of smoking and substance abuse), limited healthcare access and provisions, and the adverse effects of antipsychotic drugs (APDs) (DE Hert et al., 2011; Dickerson et al., 2016). Unfortunately, this excess medical burden among psychiatric patients is also accompanied by systemic gaps in healthcare quality which is reflected in a lack of early detection and appropriate management of somatic health concerns. Among psychiatric patients, roughly half of all physical illnesses go undiagnosed by treating physicians (Hall, Gardner, Popkin, Lecann, & Stickney, 1981; Koran et al., 1989; Koranyi, 1979). A large Danish national registry-based study found that individuals with schizophrenia were less likely to have been diagnosed with medical illnesses such as cardiovascular disease (OR = 0.55), cancer (OR = 0.38), pulmonary disease (OR = 0.42), and diabetes (OR = 0.36) even when these conditions were reported as their cause of death (Brink et al., 2019). Recent advancements in genomic sequencing and testing capabilities offer opportunities in psychiatry for modifying care and closing healthcare gaps with improved detection of genetic disorders that can inform illness trajectories and predisposition for comorbid conditions (Plummer, Gordon, & Levitt, 2016; So et al., 2020).

Certain genetic findings are likely to have relevance to the clinical course and comorbidities of psychiatric patients due to their association with multi-system syndromic diseases. In the realm of psychiatric genetics research, findings over the past two decades have partly focused on a class of structural variants known as copy-number variants (CNVs). CNVs, often defined as deletions or duplications of DNA sequences spanning > 1000 bases, contribute to overall genetic and phenotypic variability in the population (Sebat et al., 2004; Zhang, Gu, Hurles, & Lupski, 2009). There are now dozens of genetic loci at which CNVs are known to re-occur in the general population and which carry significant risk for neurodevelopmental disorders (NDD CNVs) such as schizophrenia (ORs ranging from 1.8 – 67.7) (Marshall et al., 2017). These NDD CNV sites are predisposed to recurrent de novo CNV formation (Pös et al., 2021). It is estimated that ∼ 2 – 3% of patients with schizophrenia carry one of these NDD CNVs (Marshall et al., 2017; Rees et al., 2016; Rees et al., 2014). These rates also vary depending on patients’ clinical presentation; there is an increased likelihood of being a CNV carrier among patients with a childhood onset of psychotic symptoms (Brownstein et al., 2022), comorbid neurodevelopmental disorders (Foley et al., 2020), or poor clinical response to APDs (Farrell et al., 2023; Sun et al., 2024). Of importance here, NDD CNVs also have wide-ranging medical consequences that are largely underappreciated in psychiatry.

Studies describing the syndromic presentations associated with specific CNVs have mostly focused on child and adolescent patients, often highlighting risk of congenital defects and multi-system health conditions but with little or no data on their effects throughout later stages of life (Burnside et al., 2011; Butler & Thompson, 2000; Jacquemont et al., 2011; Rodríguez-Caballero et al., 2010). Recent studies involving population based cohorts in the UK biobank comparing NDD CNV carriers and non-carriers have shown that this group of recurrent CNVs also contributes to increased medical morbidity in adult populations, but data are limited by their ‘healthy volunteer’ selection bias (Aguirre, Rivas, & Priest, 2019). Crawford et al., 2019 (Crawford et al., 2019) found that these CNVs increased risk for 45% of all medical conditions that were examined (26 / 58 conditions), with “death during follow-up” being the second most significant phenotype among CNV carriers overall. Most of these CNVs remain poorly understood in terms of their broad health outcomes and clinical relevance in adult psychiatric patients. However, there are exceptions, such as with 22q11.2 deletion syndrome (Sullivan & Owen, 2020). For 22q11.2 deletion syndrome, there are now published guidelines including safety and tolerability data regarding the use of antipsychotic and antidepressant drugs in this population (Dori, Green, Weizman, & Gothelf, 2017; Tanham, Chen, Warren, Heussler, & Scott, 2024). These guidelines provide dosing standards and practices for monitoring side effects as psychiatric medications can increase the risk for a host of common medical issues seen in 22q11.2 deletion syndrome, including cardiac arrhythmias, seizures and movement disorders (Berardelli et al., 2024; Choi et al., 2020; Mosheva, Korotkin, Gur, Weizman, & Gothelf, 2020; Tanham et al., 2024). This example may just be the leading edge of a much deeper knowledge base regarding broad and specific CNV risks and their relevance to the practice of psychiatric medicine.

It is a major challenge to accumulate the amount of data necessary for risk prediction and evidence-based management in psychiatry given the rarity of high impact mutations such as NDD CNVs and the dearth of genetic testing that takes place within adult psychiatric settings. While achieving large numbers of deeply phenotyped psychiatric cases will be an important endeavor, case reports are a valuable and inexpensive way to document novel observations in the short term using richly detailed life course data (Sullivan & Owen, 2020). Here, we report the phenotypic findings from 24 adults with treatment resistant psychosis (TRS) identified as carriers of NDD CNVs.

## Methods

### Cohort Ascertainment and Genomic Screening

We previously identified 24 cases with known NDD CNVs from a sample of 509 patients with TRS. These patients were recruited between 2016-2019 from the Pennsylvania state hospital (PASH) network. Participant selection, sample processing, and genomic analysis of this PASH TRS cohort has been previously described with full sample demographic and clinical characteristics (Farrell et al., 2023). Briefly, this cohort consists of individuals who resided for a minimum of five consecutive years in a PASH and/or affiliated community-based long-term structured residential (LTSR) and had clinically validated TRS. Criteria for validating TRS included a lack of clinical improvement despite ≥3 APD trials of adequate dose and duration, the absence of any previous sustained response to treatment, and the absence of any diagnosis causative of psychosis (i.e., substance use disorder or other medical condition). This study was approved by the Institutional Review Boards at Drexel University College of Medicine and The University of North Carolina at Chapel Hill, and the Chief Medical Officer of the Commonwealth of Pennsylvania. Research was carried out in accordance with the Declaration of Helsinki. Written informed consent was obtained from all the participants (or legal guardians, if applicable), which allowed for patient re-contact.

Genomic analysis implemented CNV calling from chromosomal microarray (CMA) and whole exome sequencing (WES) data. Separate clinical-grade Agilent comparative genome hybridization array in a CLIA-certified lab (Allele Diagnostics, Spokane WA) was also sought for 22 of 24 cases in this report and confirmed the research-grade CNV calls. The genomic findings for each case are summarized including the NDD CNVs of interest and additional (second hit) CNVs. Four of these cases have been individually described in separate case reports (Farrell et al., 2018; Farrell et al., 2020; Harner et al., 2020; Xavier et al., *under review*).

### Phenotyping

PASH participants resided for years (and sometimes decades) receiving inpatient care with on-site nursing and medical care including comprehensive annual evaluations. Phenotypic and treatment data were derived from multiple sources including extensive review of active and archival medical records and a specialized neuro-behavioral examination with the goal of achieving a complete account of lifetime phenotypic manifestations across all systems. Archival information often dated back to individuals’ school records, earliest psychiatric treatment in adolescence or early adulthood and included annual psychiatric, psychological, nursing, and medical evaluations. A subset of participants (9 of 24 cases presented here) gave consent to be evaluated by a behavioral neurologist. This consultation (by author M.L.L.) obtained; 1) family, social, developmental, treatment, and medical history, 2) a physical examination, 3) a complete neurological examination and, 4) a summary of findings in the context of their genetic findings.

Each participant was also evaluated by a trained examiner using the Positive and Negative Syndrome Scale (Kay, Fiszbein, & Opler, 1987) and the Ammons Quick Test of intellectual functioning (Ammons & Ammons, 1962). The Wechsler Adult Intelligence Scale-III (WAIS) (Wechsler & De Lemos, 1981) was administered to participants in this report who could engage in the testing (N = 9). Results from prior intelligence testing were taken from medical records when available (N = 3). Data were systematically extracted and managed using REDCap electronic data capture tools hosted at the University of North Carolina at Chapel Hill (Harris et al., 2019). Plots were generated using ComplexUpset (Krassowski, Arts, Lagger, & Max, 2022), ggforce (v0.4.2) (Pedersen, 2024), and ggplot2 (Wickham, 2016) packages in R Statistical Software (v4.4.1) (R Core Team, 2024).

The Human Phenotype Ontology (HPO) was used to code phenotypes and synthesize terminology across individuals for a standardized and reproducible presentation. Phenotypes were mapped to HPO terms or synonyms between February 2023 – July 2023 using the HPO online browser tool (Gargano et al., 2024). This mapping and synthesis often required making close approximations to data extracted from patient medical records and considerations for the evolving diagnostic clinical terminology used over the decades of care these records often covered. For example, the notion of specific learning disabilities (SLDs) in childhood was not introduced into official psychiatric nomenclature until the second edition of the Diagnostic and Statistical Manual of Mental Disorders (American Psychiatric Association, 1968). At that time, the DSM-II introduced the term *specific learning disturbance* in Special Symptoms Not Elsewhere Classified (306.1); however, no specific disabilities were delineated, nor were criteria specified beyond stating that the “psychopathology is manifested by a single specific symptom”. When reviewing archival records, terms for “single symptoms” were often found that reflected childhood deficiencies introduced much earlier by Travis, Kirk, and others, such as “disorders of attention”, “deficiencies in visual and auditory memory”, “stuttering”, “alexia”, and “aphasia” (Kirk, 1963; Travis, 1935). Where archival records were available, all SLD notations were extracted from the early educational and/or psychiatric records and tabulated across the life-span, always distinguishing between earlier “descriptive” notations and official diagnostic (DSM) codes.

HPO terms and their corresponding IDs were extracted, taking the most specific term possible given available data. The HPO terms that roughly correspond to the major organ systems (i.e., immune system, digestive system, integument, etc.) were used to categorize the specific phenotypic terms, grouping each specific phenotype to only one organ system (a simplified grouping as many specific terms actually belong to multiple parent terms in the HPO system, i.e., asthma is an abnormality of the immune system *and* is an abnormality of respiratory system). HPO terms for symptoms of psychosis (positive and negative symptoms) are not included as these cases were selected for the persistence of these symptoms. Additionally, the overall rate of drug allergies is counted, but specific iatrogenic phenotypes (surgical procedures, specific adverse drug effects) and behaviorally acquired phenotypes (head injury, sexually transmitted diseases) are discussed separately from the organ systems in which they occur. Phenotypes are examined in aggregate for the entire group (N = 24) and within sets of individuals that carry CNVs in common (22q11.2 deletions, 15q11.2 deletions, etc.).

## Results

From a cohort of patients with TRS, 24 cases were identified as carriers of known recurrent NDD CNVs (4.7% of the PASH cohort). Some demographic and NDD CNVs findings for these cases were previously presented (Farrell et. al, 2023). The mean age at study enrollment was 53.3 years (range 23 – 71 years). The most common primary Axis I diagnosis at enrollment was schizophrenia (13/24 cases, any subtype), followed by schizoaffective disorder (7/24 cases). Several CNV loci were identified in more than one case, including 15p11.2 deletions (3 cases), 15q11.2q13.1 duplications (4 cases), proximal 16p11.2 duplications (6 cases), and 22q11.2 deletions (4 cases).

CNVs with less or no established clinical significance (“second hits”) were identified within the genomes of these 24 patients. Two cases had second hit CNVs overlapping known disease-associated genes; a deletion of SLC5A7 in *case 2 (Barwick et al., 2012),* and a duplication partially overlapping PRKN in *case 13 (Portman et al., 2001; Yoshikawa et al., 2022)*. Of the additional genetic findings, one case (*case 19*) was found to carry a trinucleotide expansion in the HTT gene (48-copy CAG allele) pathogenic for Huntington’s disease following the neuro-behavioral evaluation, in addition to the 22q11.2 deletion (Farrell et al., 2018).

### Phenotypic Findings

Focusing first on somatic systems only, not including psychiatric and behavioral phenotypes, **Figure 1** summarizes; 1) organ systems affected (by one or more phenotypic manifestations) within each case, and 2) the frequency of affected systems across the cohort. Individuals had phenotypic manifestations across a median of 10.5 organ systems (range 5 – 15 affected systems per individual). This multi-system comorbidity was highest in the 7q11.23 duplication carrier (15 affected systems), followed by the group of proximal 16p11.2 duplication carriers (6 cases, range = 9 – 13 affected systems) and 15q11.2q13.1 duplication carriers (4 cases, range = 10 – 13 affected systems). The most frequently affected systems overall were cardiovascular, motor, metabolic, and digestive systems. Within each organ system, these 24 cases demonstrated a constellation of specific phenotypes which at the individual level reflects an extreme degree of medical comorbidity. For further examination, specific phenotypic findings <u>affecting two or</u> <u>more cases</u> are separated into three categories and summarized below: 1) general medical disorders, 2) neurological and motor disorders, and 3) psychiatric and developmental disorders. Importantly, numerous additional phenotypes were less common and seen in only one case.

**Figure 1.**
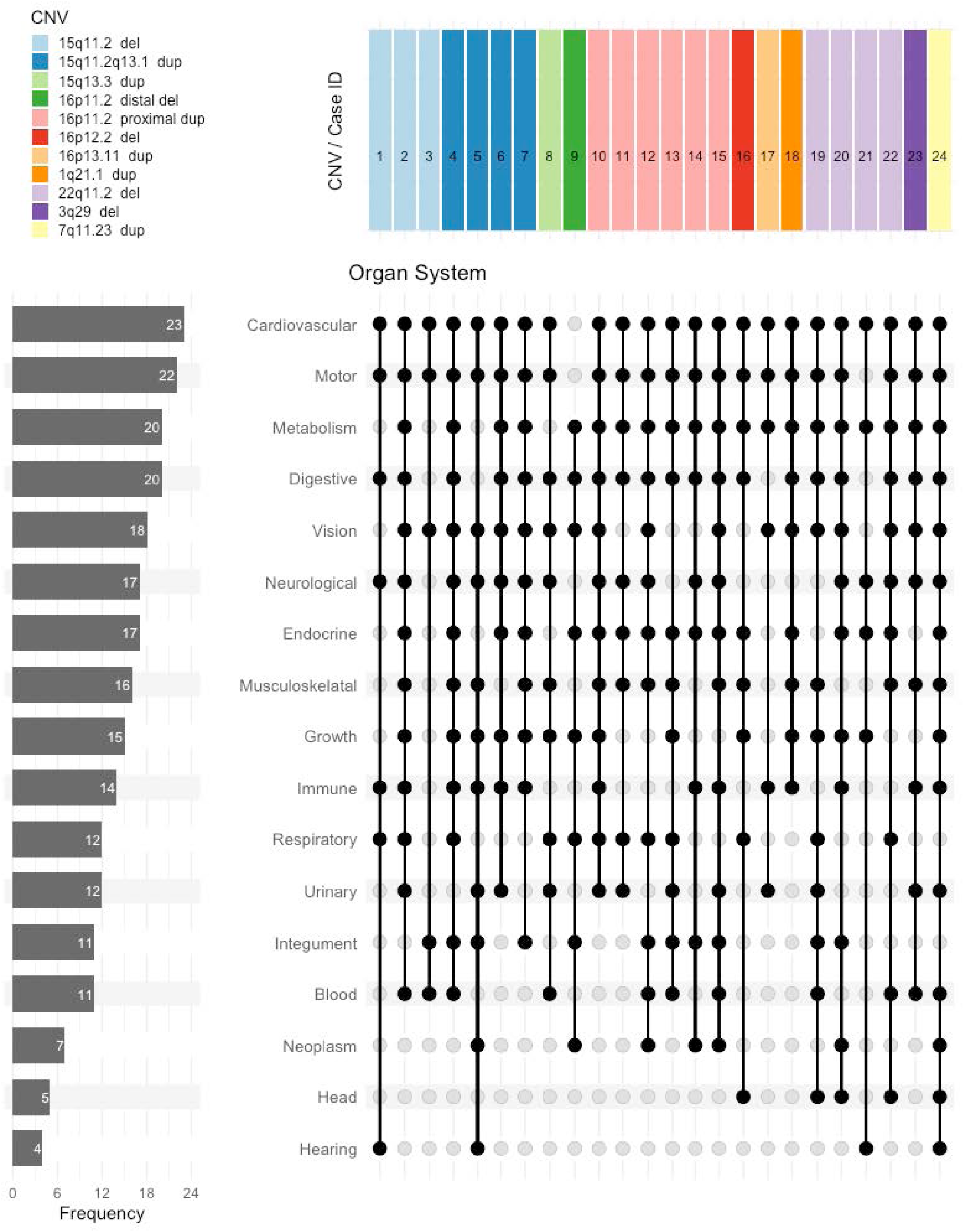
Affected organ systems by case. Affected organ systems within each case represented by filled in dots with connecting lines (vertical columns) in Upset matrix. Case IDs are shown (top) within the colored bars corresponding to the NDD CNV loci (CNV legend). Filled in dots indicate organ systems affected by at least one HPO phenotype and the horizontal bar graph (left) shows the group-level frequency of affected organ systems.

**Table 1.** Demographic and Genomic Findings.

| Demographics |  |  |  | Clinical and genetic characteristics |  |  |  |  |  |  |  |
| --- | --- | --- | --- | --- | --- | --- | --- | --- | --- | --- | --- |
| Case | Sex | Race | Ethnicity | Primary Diagnosis | Full-scale IQ estimate | IQ measure | CNV band | CNV Type | CLIA | Neurological Evaluation | Additional |
| 1 | Male | White | N-H | Scz | 76 | WAIS | 15q11.2 | Del | Yes | Yes |  |
| 2* | Female | White | N-H | Scz-Aff | 61 | WAIS | 15q11.2 | Del | Yes | Yes |  |
| 3 | Female | Black | N-H | Scz-Aff | 66 | WAIS | 15q11.2 | Del | Yes | No |  |
| 4 | Male | White | N-H | Scz-Aff | 62 | WAIS | 15q11.2q13.1 | Dup | Yes | Yes |  |
| 5 | Male | White | N-H | Scz |  |  | 15q11.2q13.1 | Dup | Yes | No |  |
| 6 | Male | White | N-H | Scz-Aff | 66 | WAIS | 15q11.2q13.1 | Dup | Yes | Yes |  |
| 7 | Female | White | N-H | Psychosis NOS | 91 | WAIS | 15q11.2q13.1 | Dup | Yes | Yes |  |
| 8 | Male | Black | N-H | Scz | 69 | WAIS | 15q13.3 | Dup | Yes | No |  |
| 9 | Female | White | N-H | Scz-Aff |  |  | 16p11.2 | Del | Yes | No |  |
| 10 | Male | White | N-H | Bipolar I w/ psychosis | 104 | Ammons | 16p11.2 | Dup | Yes | No |  |
| 11 | Male | White | N-H | Scz | 71 | WAIS* | 16p11.2 | Dup | Yes | No |  |
| 12 | Male | White | N-H | Scz-Aff |  |  | 16p11.2 | Dup | Yes | No |  |
| 13 | Male | White | N-H | Scz | 85 | Shipley<br>Hartford | 16p11.2 | Dup | Yes | No |  |
| 14 | Male | White | N-H | Psychosis NOS |  |  | 16p11.2 | Dup | Yes | No |  |
| 15 | Female | White | N-H | Major Depressive<br>Disorder w/ Psychosis | 100 | Ammons | 16p11.2 | Dup | Yes | No |  |
| 16 | Male | White | N-H | Scz |  |  | 16p12.2 | Del | Yes | No |  |
| 17 | Female | White | N-H | Scz | 87 | WAIS* | 16p13.11 | Dup | Yes | No |  |
| 18* | Male | Black | N-H | Scz | 92 | Ammons | 1q21.1 | Dup | No | No |  |
| 19* | Male | White | N-H | Scz |  |  | 22q11.2 | Del | Yes | Yes | Huntington's Disease |
| 20 | Female | White | H | Scz | 92 | Ammons | 22q11.2 | Del | Yes | Yes |  |
| 21 | Female | White | N-H | Scz |  |  | 22q11.2 | Del | Yes | No |  |
| 22 | Male | White | N-H | Scz |  |  | 22q11.2 | Del | Yes | Yes |  |
| 23* | Female | White | N-H | Scz | 62 | WAIS | 3q29 | Del | Yes | Yes |  |
| 24 | Male | White | N-H | Scz-Aff | 87 | Ammons | 7q11.23 | Dup | No | No |  |

Cases with an \* have been previously reported (citations in main text). N-H = non-hispanic, H = hispanic ancestry. Scz = schizophrenia diagnosis, any subtype, Scz-aff = sczhizoaffective disorder diagnosis at time of study enrollment. WAIS\* = scores from previously administered intelligence testing taken from medical chart. Del = deletion, Dup = duplication. CLIA = CNV confirmed by an independent clinical grade (CLIA-certified) lab.

### General Medical Phenotypes

All general medical phenotypes <u>affecting two or more cases</u> are summarized in **Figure 2**. Across these systems, 14 specific medical phenotypes were present in six or more cases (≥25% of the cohort). Individuals experienced a median of 11 distinct medical phenotypes (range 4 – 22). The number of distinct general medical phenotypes per individual was notably greatest in the one 7q11.23 duplication carrier (22 phenotypes), again followed by the group of proximal 16p11.2 duplication carriers (6 cases, range 12 – 19 phenotypes).

**Figure 2.**
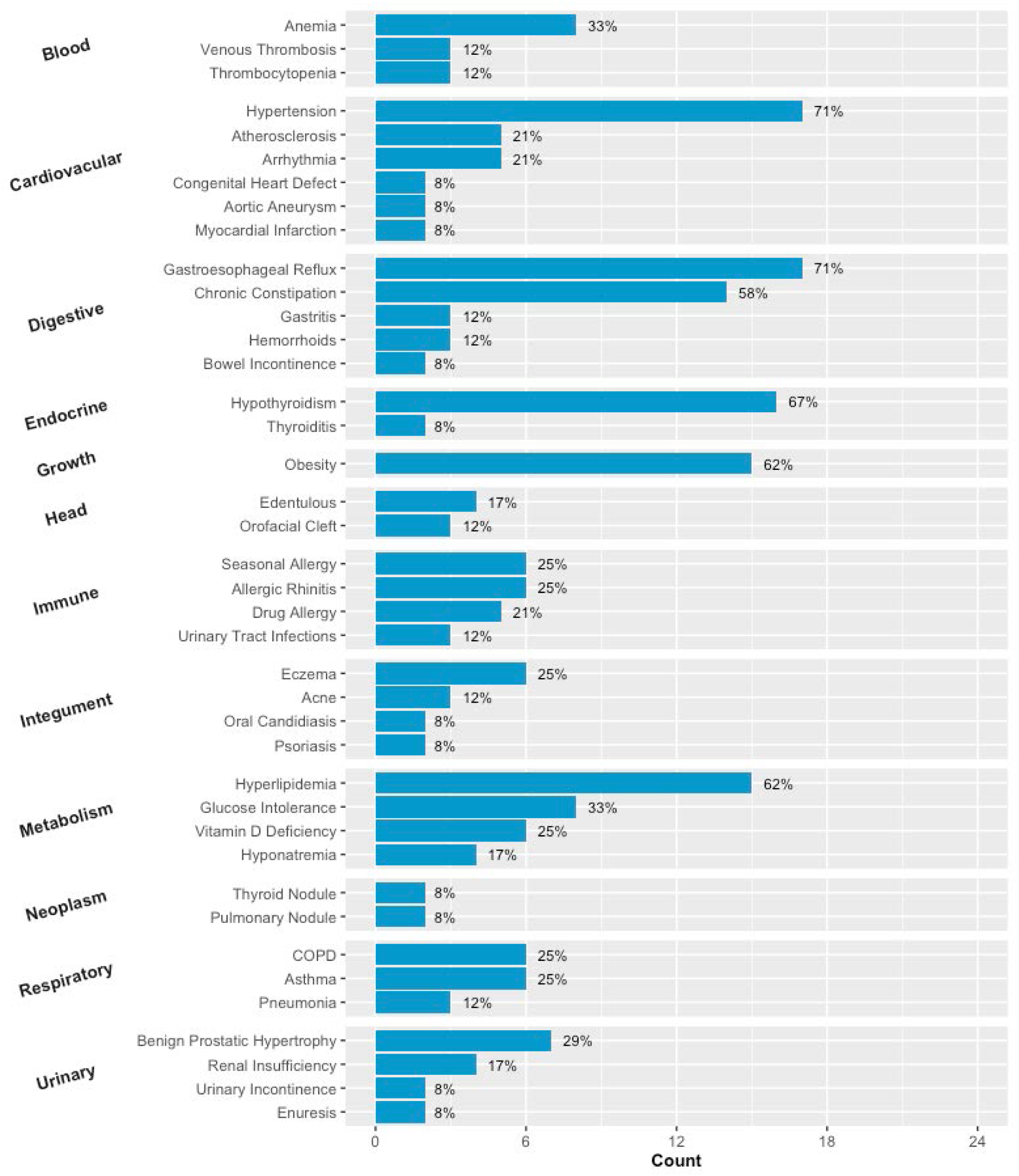
General medical phenotypes. General medical phenotypes documented throughout the life course of CNV carriers, organized by organ system. Only phenotypes present in two or more cases shown here. The complete list of specific phenotypes, individual-level data, and Human Phenotype Ontology IDs are in supplemental table S2. COPD = chronic obstructive pulmonary disease.

### Other Neurological, Sensory, Musculoskeletal, and Motor Phenotypes

Phenotypes affecting motor, musculoskeletal, neurological, and sensory organ systems are examined together in **Figure 3**, again showing only phenotypes affecting two or more cases. Across these systems, nine separate phenotypes were present in six or more cases (≥25%). Individual cases experienced a median of 7 distinct phenotypes (range 1 – 10). The most prominently affected cases included the 3q29 deletion carrier (10 phenotypes), the 7q11.23 duplication carrier (9 phenotypes), and the proximal 16p11.2 duplication carriers having a group median of 8 phenotypes (6 cases, range 2 – 9 distinct phenotypes).

**Figure 3.**
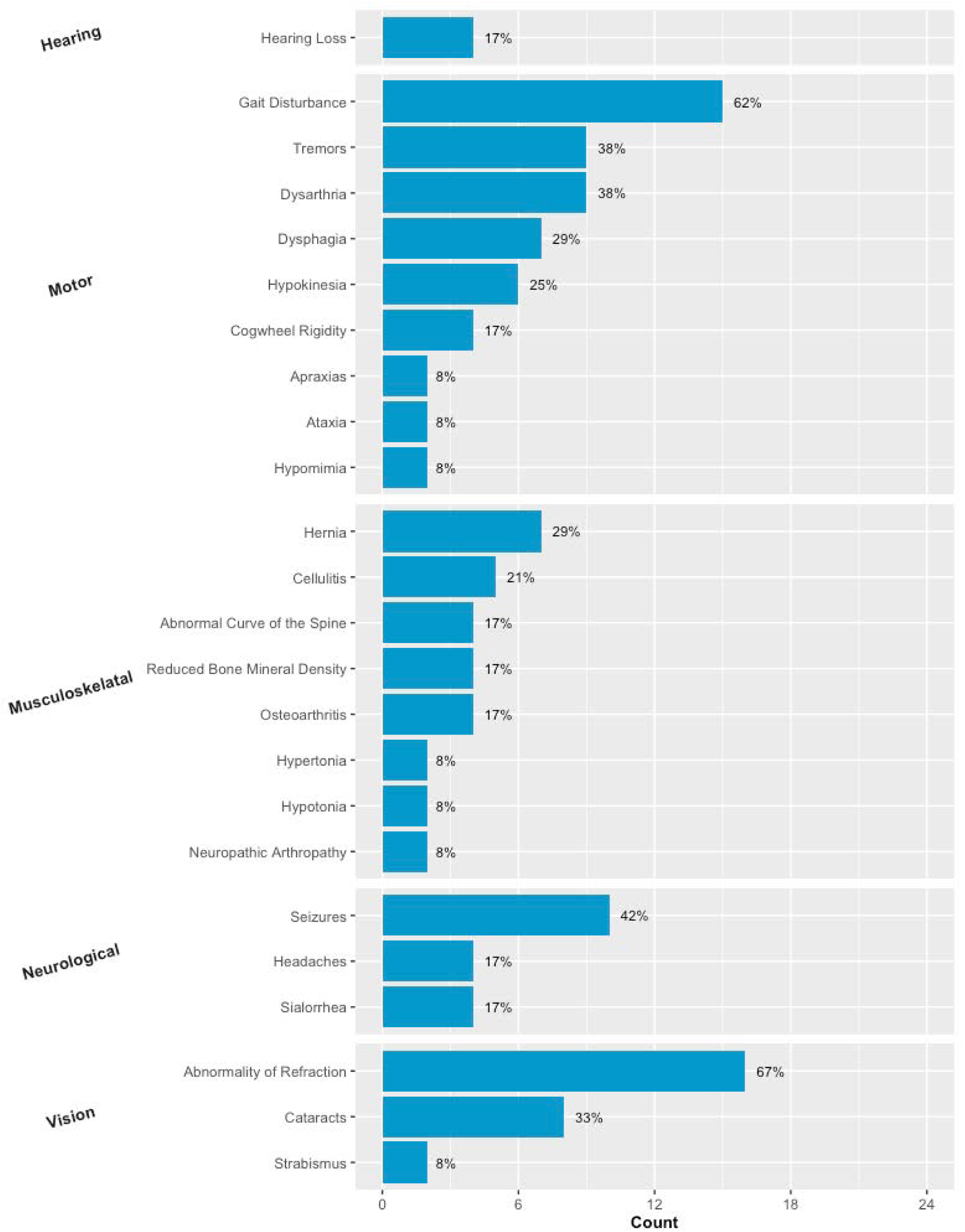
Neurological, motor, musculoskeletal, and sensory phenotypes. Phenotypes across neurological, motor, musculoskeletal, and sensory organ systems as documented throughout the life course of CNV carriers or upon neurological examination. Only phenotypes present in two or more cases shown here. See supplemental table S2 for complete individual-level data.

### Psychiatric and Neurodevelopmental Phenotypes

Psychiatric, neurocognitive, and developmental phenotypes are shown in **Figure 4**. Phenotypes affecting only one case or related to core symptoms of psychosis (positive and negative symptoms) are not shown here. Across these systems, 12 separate phenotypes were present in six or more cases (≥25%). Individual cases experienced a median of 10 distinct phenotypes (range 3 – 16). The most prominently affected cases included the 15q11.2 deletion carriers with a range of 7 – 15 phenotypes, the 3q29 deletion carrier (11 phenotypes), the 15q13.3 duplication carrier (11 phenotypes), and the group of 15q11.2q13.1 duplication carriers with a range of 10 – 16 phenotypes.

### CNV-specific observations

Several phenotypes appeared to be concentrated within sets of individuals carrying CNVs in common. For example, all three 15q11.2 deletion carriers had developmental speech delays and were characterized as having autistic symptoms and a happy demeanor. All four 15q11.2q13.1 duplication carriers had anxiety, ADHD symptoms, and self-injurious behaviors. Within the four 15q11.2q13.1 duplication carriers, two had diminished motor strength upon neurological examination (hypotonia), and disorders affecting the visual system (cortical visual impairment, retinal drusen, and optic nerve atrophy, strabismus, cataracts, refractive errors) were present in all four 15q11.2q13.1 duplication cases. Obesity was also observed in all four 15q11.2q13.1 duplication carriers compared to only two (out of six) 16p11.2 proximal duplication carriers. This is consistent with known associations of 16p11.2 duplications and decreased BMI (D’Angelo et al., 2016).

Within the six proximal 16p11.2 duplication carriers, glucose intolerance (prediabetes and diabetes) and cardiac arrhythmias were observed in three cases each. Several 16p11.2 duplication carriers experienced either dysphagia (3 cases), dysarthria (2 cases), or sialorrhea (drooling, 2 cases), broadly implicating the oral-facial neuromuscular system. Five 16p11.2 duplication carriers experienced chronic speech difficulties involving soft/mumbled, slowed, or slurred speech production (most not diagnosed with dysarthria). The majority of 16p11.2 duplication carriers also had diminished motor activity (hypokinesia, 4 cases). Four 16p11.2 duplication carriers had either dementia (3 cases)or mild cognitive impairments (1 case). All three 16p11.2 duplication carriers with dementia also had abnormal consumption behaviors; one case had pica and psychogenic polydipsia, and two experienced restrictive eating behaviors during catatonic episodes requiring assistance with feeding. The two proximal 16p11.2 cases with dementia and restrictive eating also presented with mutism/severe alogia during catatonic episodes.

Cases with congenital defects were all carriers of CNVs for which such morphological changes are a known risk; congenital heart defects were seen in one 15q11.2q13.1 duplication carrier and one 22q11.2 deletion carrier, and orofacial cleft in three (out of four) 22q11.2 deletion carriers. Other cardiac phenotypes (heart murmurs, arrythmias, hypertension) were also common in the subset of 22q11.2 deletion carriers. Furthermore, anxiety and speech difficulties were each seen in three (out of four) 22q11.2 deletion carriers, Speech problems in these cases were characterized as aphasia in one case (with Huntington’s disease) and dysarthria/slurred speech in two cases. Huntington’s disease itself was not “counted” as a separate phenotype for this individual but does contribute to their overall clinical presentation.

### Psychotropic Treatment and Antipsychotic Drug Side Effects

The most prescribed APDs in the group were haloperidol (21/24), olanzapine (19/24), risperidone (18/24), and chlorpromazine (17/24). Eleven (of the 24) cases were tried on clozapine and no serious adverse events were reported. Nineteen (of the 24) cases had treatment histories in a PASH with available electronic prescription records. From these, we calculated the average daily dose of all APD prescriptions (converted to CPZ equivalents) over the course of their time in the state hospital system. The median average daily dose among the group was 874.6 mg CPZ (range 192.3 – 2940.7 mg CPZ). This was highest among the 16p11.2 duplication carriers with a group median of 975.2 mg CPZ (range 475 – 2940.7 mg CPZ). Nearly all cases were also treated with mood stabilizers (21/24 cases) including lithium for 11 cases, antidepressants (20/24 cases), and anxiolytics (21/24 cases).

With archival records and annual evaluations / summaries being a primary source, there was a limited ability to discern various health changes across time (weight gain, movement symptoms) as being directly related to treatment side effects unless specific diagnoses were given (such as tardive dyskinesia). Such side effects and other specific iatrogenic conditions were not included as phenotypes in the above sections or figures but were present in several cases. The most common treatment side effect was tardive dyskinesia (N = 7), including in two (out of three) 15q11.2 deletion carriers. The one 15q13.3 duplication carrier (*case 8*) had extrapyramidal symptoms not otherwise specified. Neuroleptic malignant syndrome occurred in one 16p11.2 duplication carrier (*case 12*). Oculogyric crisis occurred in one case (*case 17*).

## Discussion

There is a pressing need to address multimorbidity and the specific health challenges that pose undue stress and economic burden, and contribute to increased suffering and mortality in patients with serious mental illness. The strongest genetic risk factors currently known to increase risk for neuropsychiatric disorders, NDD CNVs, are also known to increase general medical burden. The patients described here illustrate a complex course of illness, complicated by medical and neuropsychiatric multimorbidity. While the influence of the NDD CNVs in these cases likely contribute to this morbidity, the clinical interpretation of even highly pathogenic CNVs is complicated by their notoriously variable phenotypic expression and incomplete penetrance for any specific health outcomes (Nowakowska, 2017). Thus, the complexity of these cases should be appreciated, first, even irrespective of their genetic findings. Potential CNV effects are also challenging to disentangle in these cases due to the influences of other factors such as chronic APD exposure, prolonged institutionalization, and age-related illness. Yet, observations from these cases can be interpreted with caution to unveil new insights on the longitudinal effects of these variants.

When examining the OMIM and ORPHANET linked phenotypes for the NDD CNVs discussed here, it is not surprising that the majority (∼75%) of phenotypes fall within the broad categories of developmental defects, dysmorphic features, and psychiatric / behavioral disturbances (Rubinstein, Church, & Maglott, 2015). This early-life bias is a strong reflection of the pediatric populations in which CNVs have been preferentially detected and reported. 22q11.2 deletion syndrome is an outlier in this regard and comparatively has a more robust database with known phenotypes spanning numerous somatic organ systems. This wider representation is derived from collaborative efforts, the systematic inclusion of many adult cases, and the collation of medical guidelines from multi-disciplinary perspectives (Boot et al., 2023). Thus, expectedly, many of the observations from the 22q11.2 deletion carriers presented here agree with already known health risks of this CNV (GERD, thrombocytopenia, hypothyroidism, obesity, etc.). While the practice of psychiatry is still in its infancy as far as extracting clinical information from the rapidly advancing field of genetics, the 22q11.2 deletion syndrome is one of the few variants with such a robust literature and potential to inform treatment decisions.

In contrast to 22q11.2 deletion syndrome, there is far less concordance between clinical presentations of other CNV carriers presented here (15q11.2 deletions, 15q11.2q13.1 duplications, etc.) and their known health risks. Novel observations from these cases may prove valuable for clinicians and future research. For example, there are no prior data supporting the likelihood of several phenotypes that were prominent among the six 16p11.2 duplication carriers, including catatonia (2/6 cases), restrictive eating behaviors (2/6), arrhythmias (3/6 cases), atherosclerosis (3/6 cases), hypokinesia (4/6 cases), dementia or mild cognitive impairment (4/6 cases, early onset in 2 cases), and speech disturbances (mutism, bradylalia, soft, or slurred speech; 6/6 cases). Similarly, all four 15q11.2q13.1 duplication carriers suffered from problems of their visual systems (retinal drusen, optic nerve atrophy, cortical vision impairment, strabismus, cataracts, refractive errors). The full set of phenotypes could be leveraged in further meta-analytic studies to elucidate such risks more precisely from detailed individual-level data (Sullivan & Owen, 2020).

In conclusion, this opportunity to explore comprehensive life-course data for genotyped adults from psychiatric settings demonstrates both group-level and potential CNV loci-specific insights into the complex phenotypic manifestation of these variants. Psychiatric care is often compartmentalized from other medical disciplines despite the impact of burdensome medical comorbidities. Understanding the full scope and clinical implications of recurrent genetic variants may offer opportunities for better integration of care, and more precise and individualized risk detection. Despite the surplus of archival records that were available for many of these cases, this analysis was retrospective in nature, and thus details of early development including precise measurement of more subtle dysmorphic features, developmental milestones, and other measurable phenotypes are lacking. A specialized neurobehavioral evaluation was also available for some cases (N = 9), but not all. Furthermore, the standard practice for case reports to provide data on specific clinical outcomes including the efficacy of care strategies are also limited for these cases. This is especially true of their psychiatric care given the *a priori* selection for APD treatment resistance. Despite these limitations, these observations remain relevant to the current challenge of disentangling the multimorbidity associated with rare CNVs in the context of psychiatric care.

## Data Availability

All data produced in the present study are available upon reasonable request to the authors.

## Acknowledgements

The authors thank Dale Adair, MD, Deputy Secretary of Health and Human Services, and Chief Medical Officer, Office of Mental Health and Substance Abuse Services, Commonwealth of Pennsylvania, for his encouragement and support for this project. The authors also thank Marice Davis for providing provided phlebotomy services. We also thank the NIMH and Rutgers University Cell and DNA Repository for support in sample collection and processing. PFS reports financial interests from H. Lundbeck A/S (advisory committee), the H. Lundbeck Foundation (grant recipient), and RBNC Therapeutics (advisory committee). Some of the sample recruitment and genomic assays were supported by H. Lundbeck A/S.

## Funding

This work was supported by the National Institute of Mental Health (K01 MH108894, R01MH110427 and K99HG008689); H. Lundbeck A/S; the Vernik Family Trust; and the Samuel and Paul Lofgren Family Trust.

## Notes

### Competing Interest Statement

The authors have declared no competing interest.

### Funding Statement

This study did not receive any funding.

### Author Declarations

Approval for this project was granted by the IRBs at Drexel University College of Medicine and the University of North Carolina at Chapel Hill, as well as the Chief Medical Officer of the Department of Health and Human Services of the Commonwealth of Pennsylvania

